# Existing human mobility data sources poorly predicted the spatial spread of SARS-CoV-2 in Madagascar

**DOI:** 10.1101/2021.07.30.21261392

**Authors:** Tanjona Ramiadantsoa, C. Jessica E. Metcalf, Antso Hasina Raherinandrasana, Santatra Randrianarisoa, Benjamin L. Rice, Amy Wesolowski, Fidiniaina Mamy Randriatsarafara, Fidisoa Rasambainarivo

## Abstract

For emerging epidemics such as the COVID-19 pandemic, quantifying travel is a key component of developing accurate predictive models of disease spread to inform public health planning. However, in many LMICs, traditional data sets on travel such as commuting surveys as well as non-traditional sources such as mobile phone data are lacking, or, where available, have only rarely been leveraged by the public health community. Evaluating the accuracy of available data to measure transmission-relevant travel may be further hampered by limited reporting of suspected and laboratory confirmed infections. Here, we leverage case data collected as part of a COVID-19 dashboard collated via daily reports from the Malagasy authorities on reported cases of SARS-CoV-2 across the 22 regions of Madagascar. We compare the order of the timing of when cases were reported with predictions from a SARS-CoV-2 metapopulation model of Madagascar informed using various measures of connectivity including a gravity model based on different measures of distance, Internal Migration Flow data, and mobile phone data. Overall, the models based on mobile phone connectivity and the gravity-based on Euclidean distance best predicted the observed spread. The ranks of the regions most remote from the capital were more difficult to predict but interestingly, regions where the mobile phone connectivity model was more accurate differed from those where the gravity model was most accurate. This suggests that there may be additional features of mobility or connectivity that were consistently underestimated using all approaches, but are epidemiologically relevant. This work highlights the importance of data availability and strengthening collaboration among different institutions with access to critical data - models are only as good as the data that they use, so building towards effective data-sharing pipelines is essential.

## Introduction

Human mobility underlies the spatial patterns of many infectious diseases (Findlater and Bogoch, 2018; Grenfell et al., 2001; Kramer et al., 2016; Meloni et al., 2011; Tatem et al., 2006; Tizzoni et al., 2014; Wesolowski et al., 2016; Zhou et al., 2020) and will drive the dynamics of emerging epidemics. Quantifying travel patterns is key to predicting where and when the pathogen may spread and therefore to devising measures and policies to contain the epidemics. As demonstrated by the COVID-19 pandemic (Badr et al., 2020; Chang et al., 2021; Kraemer et al., 2020; Nouvellet et al., 2021), a broad range of travel from international trips to local commuting patterns drives the spatial spread of SARS-CoV-2. While data is increasingly being used to inform mobility patterns and inform predictive transmission models for public health planning (Grantz et al., 2020; Kishore et al., 2020; Oliver et al., 2020), these data are often limited in low- and middle-income countries (Gupta et al., 2020) where routine data collection of mobility patterns may be sparse (Wesolowski et al., 2016, 2015a).

Extrapolating generic patterns of mobility derived from data from High Income Countries (HIC) to Low & Middle Income Countries (LMICs) may be misleading (Wesolowski et al., 2016, 2015a) given greater subnational heterogeneity and less developed infrastructure. For instance in Madagascar, beyond the road infrastructure, which is sparse and may be in poor condition (Fig. 1A), there is one semi-functional railroad, and a handful of commercial flights directed mostly to tourist destinations. As a result, moving not only takes time, but is also highly dependent on local topography and road conditions. Although mobility data has the potential to shed light into how these limitations translate to realized mobility, high quality data on mobility are limited. There is no systematic digitized data for travelers, a small proportion of the population (41%) has access to mobile phones (Mobile cellular subscriptions), and mobility data derived from the latter are not readily available. Yet the problem of understanding spatial spread of infectious diseases is persistent and there is a need to use data to inform decision-making around resource allocation.

**Figure 1:**
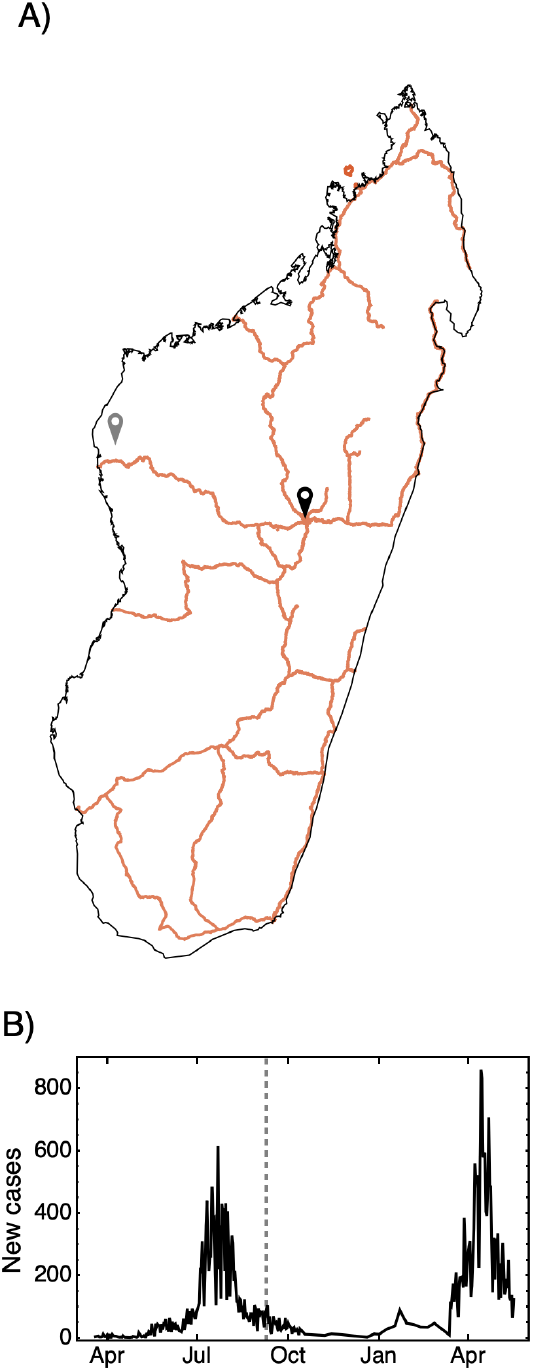
The geographic context and time-course of SARS-CoV-2 cases in Madagascar. A) The primary and secondary road infrastructure in Madagascar with the location of the first reported case (national capital of Antananarivo (black pin) of the Analamanga Region) and last region to report a confirmed case (Maintirano (grey pin) of the Melaky region). B) The confirmed cases of SARS-CoV-2 as collated on the COVID-19 dashboard (covid19mg.org) from the 20th of March 2020 to the 17th of May 2021. The date when the first case was reported in Maintirano, Melaky is shown as a grey dashed line (9th September 2020).

Madagascar is a large island 400km east of Africa with a population size of about 26 million (INSTAT-RGPH3) of which 78% earns less than 1.9 US dollar per day (UNDP-Multidimensional Poverty Index). The pandemic virus, SARS-CoV-2 was officially reported on the 20th of March 2020, with three imported cases from France arriving at the capital airport. One specific case was investigated after a tourist tested positive upon return in France prompting the first contact tracing efforts in Madagascar. About six months after the first official case, Melaky was the last of the 22 regions to have reported at least one confirmed case. Madagascar has undergone two waves with the second declared roughly one year after the first confirmed case (Fig. 1B). At the time of writing (May 2021) a total of 40,474 cases have been reported. Because all airports were closed, minimizing the risks of importation, Madagascar is a near ‘closed’ system, making it an ideal setting in which to investigate the role of internal mobility in the spread of SARS-CoV-2.

Previous work in Madagascar has noted delayed introduction and low case detection rates as important factors in shaping low numbers reported (Evans et al., 2020). We extend this work here by providing an analysis of the spatial spread of COVID-19 using various measures of mobility to identify which source of mobility is best able to reproduce the spatial dynamics of the outbreak. Lack of an official compiled and accessible database on COVID-19 in Madagascar prompted us to develop a Madagascar-specific dashboard (covid19mg.org), which is used throughout our analysis (see below). We leverage a range of data and modeling tools to better understand the spread of SARS-CoV-2 in a data limited settings reflecting highly heterogeneous demographics, accessibility, and road networks throughout the country.

## Material and methods

SARS-CoV-2 Confirmed Case Data in Madagascar: Since there is no accessible national SARS-CoV-2 database, we compiled data communicated by the Ministry of Health of the Government of Madagascar on national television every day. These include the number of new cases (confirmed by PCR), severe clinical cases, deaths, recovered, and the number of tests (data accessible at: covid19mg.org). The detail and consistency of reporting for each category varies, however the number of cases per region was reported throughout the time period allowing us to reconstruct the spatial spread across the country. The reported cases used here only include those confirmed by detection of viral nucleic acids (RT-PCR tests and or geneXpert (Rakotosamimanana et al., 2020)). Due to reporting delays, testing delays, and variable patterns of healthcare seeking behavior, we focused on the order in which cases were reported in each region. Each region was ranked based on the date when the first and fifth cases were confirmed.

In the transmission model described below, we then used various mobility matrices to model the occurrence of the first and fifth cases to compare to the reported rank.

### Mobility matrices

A mobility matrix describes how many individuals move from within and between regions per unit of time (Grenfell and Harwood, 1997). Since our goal is to understand how cases spread across regions, we ignored mobility within a region. We used four types of mobility matrices. The first three matrices are based on the gravity model with various measures of distance (Erlander and Stewart, 1990). The connectivity between region *i* and *j* is defined as 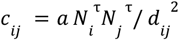, where α and *τ* are scaling factors, *N* is population size and *d*_*ij*_ represents the distance between region *i* and *j*. The distance is either the Euclidean distance (referred to as Euclidean model) between the centroids of the two regions or average transit time between the regions (referred to as transit model). To estimate the average travel time between regions (excluding flying as it is not the primary mode of transportation), we interviewed national bus companies on the travel times between the capital (Antananarivo) and the capitals of each of the 21 remaining regions. Since Antananarivo is the primary hub of travel (Fig. 1A), we calculated trips between other regions by adding or subtracting the travel times to and from Antananarivo. Travel times were directly obtained for routes that do not pass through the capital (e.g., neighboring remote regions). We also varied the parameter *τ* (0.5, 1, and 1.5) giving a total of eight mobility matrices explored.

A third gravity-based model was used. The mobility matrix is the Internal Migration Flow (flow for short) accessed from the WorldPop project (worldpop.org). The Internal Migration Flow data was developed to study the spread of malaria where no migration data is available. In short, the model estimates the number of people moving between regions by fitting a gravity model extended to account for geographic and socioeconomic factors between 2005 and 2010 (more details in (Garcia et al. 2015; Sorichetta et al., 2016)).

The fourth matrix comes from mobile phone data from Orange Madagascar, one of three main mobile phone operators in Madagascar, which records mobility traced by cell towers. Since current data is not available, we used data from a malaria study in 2015 (Ihantamalala et al., 2018).

To enable comparison among the different matrices, we standardized them so the proportion of individuals leaving each region, are on the diagonal, which we scaled so that for each mobility matrix it summed to one Departing individuals are then distributed to the other regions according to the proportions given in each column, which was scaled to sum to one but now with the diagonal element removed in order to ignore mobility within regions.The advantage of the standardization is that we can specify the total number of individuals moving per unit of time so that the mobility matrix only controls how those individuals are distributed across the regions. The technical details of the computations are reported in the supplement.

### Mechanistic model

We developed a stochastic discrete time SEIR metapopulation model for the 22 regions in Madagascar. The deterministic skeleton of the model without mobility is,

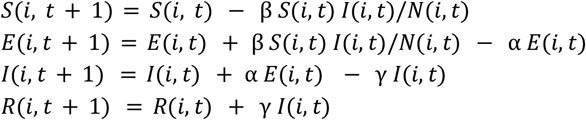

where *i* and *t* denote a region and time (days). *N(i,t)* is the population size in each region which was obtained from the National Institute of Statistics in Madagascar (INSTAT). Stochasticity is captured by setting the distribution of the number of new exposed individuals to follow a Negative Binomial(1, 1/(1+ m)) where *m =* β *S(i, t) I(i, t) / N(i, t);* similarly, the number of new infectious and new recovered individuals are drawn from a Binomial(E, α) and Binomial(I, γ) distribution, respectively. The parameters β, α, and γ are 0.24, 1/3.6,and 1/3.4 and are the same for all the regions (Bar-On et al., 2020, MIDAS Network COVID-19).

At each time step, we draw a random sample of S, E, I, and R individuals to move from region i to region j according to their respective frequency in that population, i.e., mobility is independent of whether individuals are susceptible, exposed, infected or recovered. We first specify the total number of individuals moving which are then randomly distributed across the regions using a multinomial distribution with parameters from the mobility matrix (see section above). We simulated the model until time T= 700 days and 100 replicates. Our analysis does not depend on the magnitude of time-step chosen for the simulation nor the total number of individuals moving (set to10000), as we are comparing relative, rather than absolute arrivals in each region.

### Comparing reported and simulated data

For each simulation using Analamanga region (capital) as the initial infected location, we ranked each region based on when the first and fifth cases occurred. We then compared the empirical and simulated rankings using both the cardinality of the matched rankings and Spearman rank correlations. Finally, we explored which regions were difficult to predict using the simulations using the root mean square error (rmse) of the simulated and reportedl rank.

### Statistical model

In addition to the mechanistic model, we analyzed the statistical relationships between the mobility matrices and the order of arrival. We used the Network-Based Diffusion Analysis (NBDA) in the R statistical environment using the NBDA package v0.7.10 58. In network based diffusion analysis, the order in the regions (nodes) reported the first or the fifth case (acquire a trait) is compared to their position in the network to assess whether the trait is acquired through interactions with other nodes (Hoppitt et al., 2010). The model fits a diffusion model to the reported data, more precisely it estimates a scalar (s) that controls the importance of the diffusion matrix (here the mobility matrix) to explain the order of acquisition. Significance is obtained by comparing the log-likelihood ratio between the fitted model and a null model where the scalar is set to 0.

The data compilation, metapopulation models, and figures were conducted with Mathematica 12.0 (Wolfram Research Inc.). All code is available in the repository https://github.com/ramiadantsoa/mobilityMada.

## Results

The timing of arrival of the 1^st^ case and the 5^th^ case yielded different rankings (Fig. 2). For instance, although the Menabe (ME) and Diana (DI) regions had their first confirmed case in March, the 5^th^ case only occurred in July. Atsimo Atsinanana (AA) had less than five cases as of February 2021. Among the first five regions, the 1^st^ and 5^th^ case agree in three regions: Analamanga (AN), Atsinanana (AT), and Matsiatra Ambony (MA). For the 1^st^ case metric, the remaining regions were Menabe (ME) in the west and Diana (DI) in north whereas for the 5^th^ case metric, the remaining regions were Alaotra Mangoro (AL) in the east and Atsimo Andrefana (AD) and Anosy (AS) in the south.

**Figure 2:**
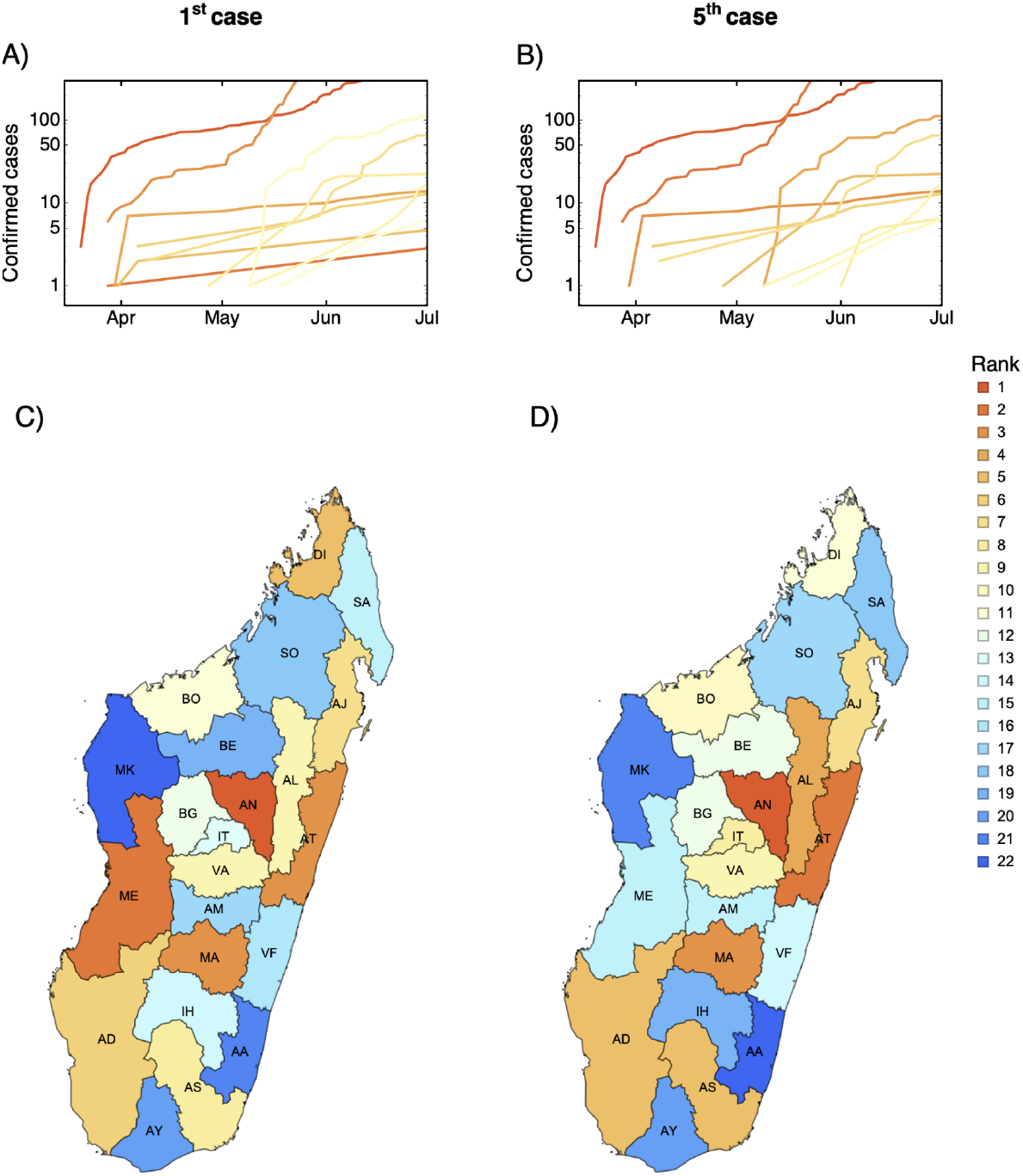
Spatial and temporal dynamic of SARS-COV-2 across the 22 regions in Madagascar. In A) and B) the time-series of the cumulative number of cases for the regions based on the first or fifth reported cases with colors corresponding to the rank. In C) and D) the distribution of the order of arrival ranked based on the first and fifth reported cases, respectively, is mapped by region. Codes: AA = Atsimo Atsinanana, AD = Atsimo Andrefana, AJ = Analanjirofo, AL = Alaotra Mangoro, AM = Amoron’i Mania, AN = Analamanga, AT = Atsinanana, AS = Anosy, AY = Androy, BE = Betsiboka, BG = Bongolava, BO = Boeny, DI = Diana, IT = Itasy, MA = Matsiatra Ambony, ME = Menabe, MK = Melaky, IH = Ihorombe, SA = SAVA, SO = Sofia, VA = Vakinankaratra, VF = Vatovavy-Fitovinany.

To assess the differences among the mobility matrices, we ranked the regions according to the number of individuals entering each region (Fig. 3). The gravity models are quite similar, the Spearman correlation between the ranks are 0.98 between the Euclidean and the Internal Migration Flow and 0.88 between the Euclidean and the transit models. The gravity model based on Euclidean distance ranks the eastern regions in the central highland higher whereas the transit model ranks the southern regions higher (Fig. 3AB). Although the east is indeed closer, the terrain is steep and windy, lengthening trip duration. The Internal Migration Flow provides a similar ranking except that it ranks Atsinanana (AT) higher, which is the second largest economic region in Madagascar, and also includes Atsimo-Andrefana (Fig. 3C). The mobile phone model is markedly different and is heterogeneous - the correlation with the Euclidean based gravity model is 0.26. The model ranks more highly the remote northern and southern regions Diana (DI) and Androy (AY) (Fig. 3D).

**Figure 3:**
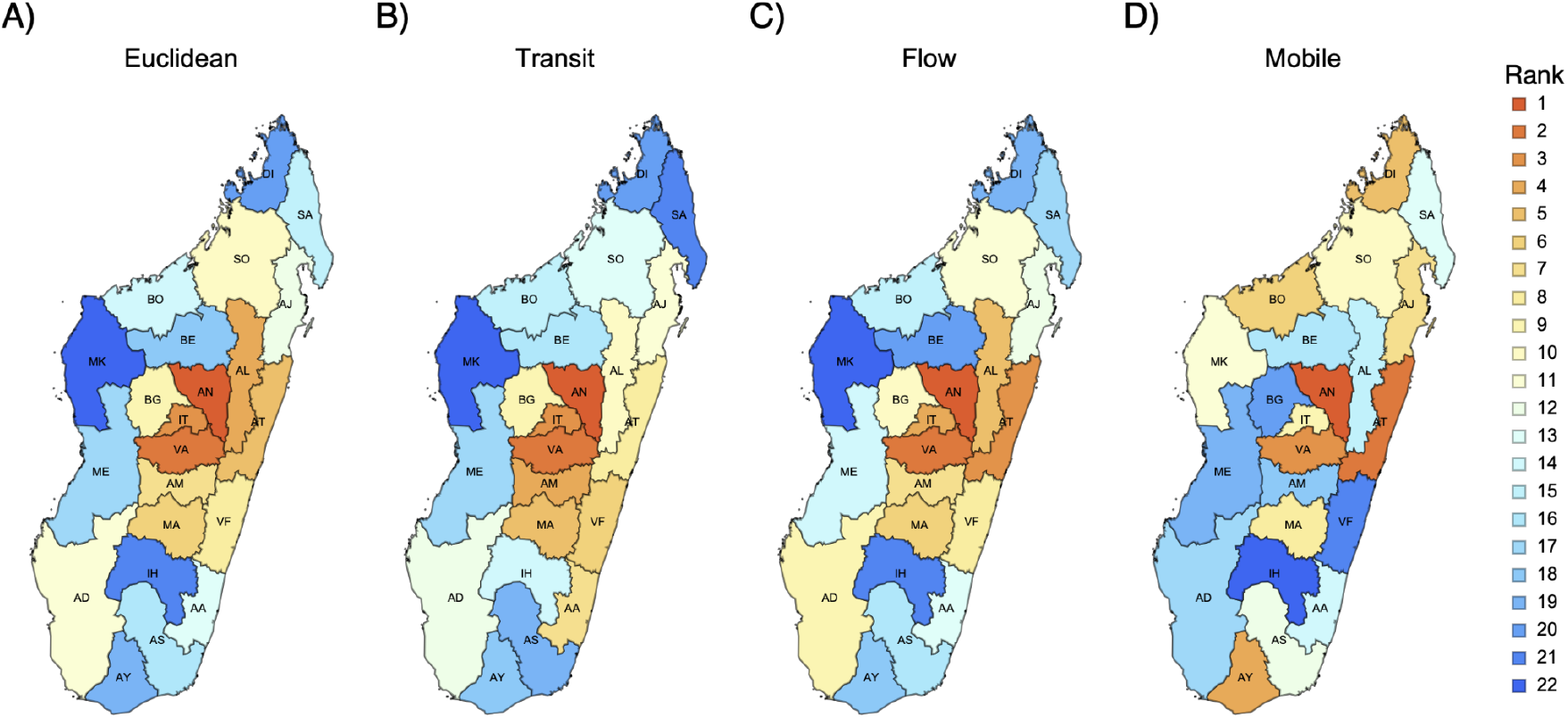
A comparison of the four main mobility matrices used. The regions are ranked according to the number of individuals entering the region per day using the various mobility matrices: A) Euclidean, B) transit, C) Internal Migration Flow (Flow), and D) mobile phone data.

Whether we looked at the overlap of the first five regions or the Spearman rank correlation for all regions, all mobility matrices better predicted the 5^th^ case than the 1^st^ case (Fig. 4 and Fig. S2). When predicting the first five regions for the 5^th^ case, the mobile phone model performed best (mean = 1.9 regions corrected predicted, Fig. 4A). Surprisingly the null model has a higher mean number of regions correctly predicted than the gravity based mobility when predicting the first five regions for the 1^st^ case (1.5 vs 1.0, 1.0, 0.9 for the Euclidean distance, transit time, and Internal Migration Flow). The Internal Migration Flow model had the worst performance among the mobility matrices investigatedFig. 4A). We also looked at the first ten regions and the results are quite similar except that the gravity models performed better than the null-model (Fig. S2).

**Figure 4:**
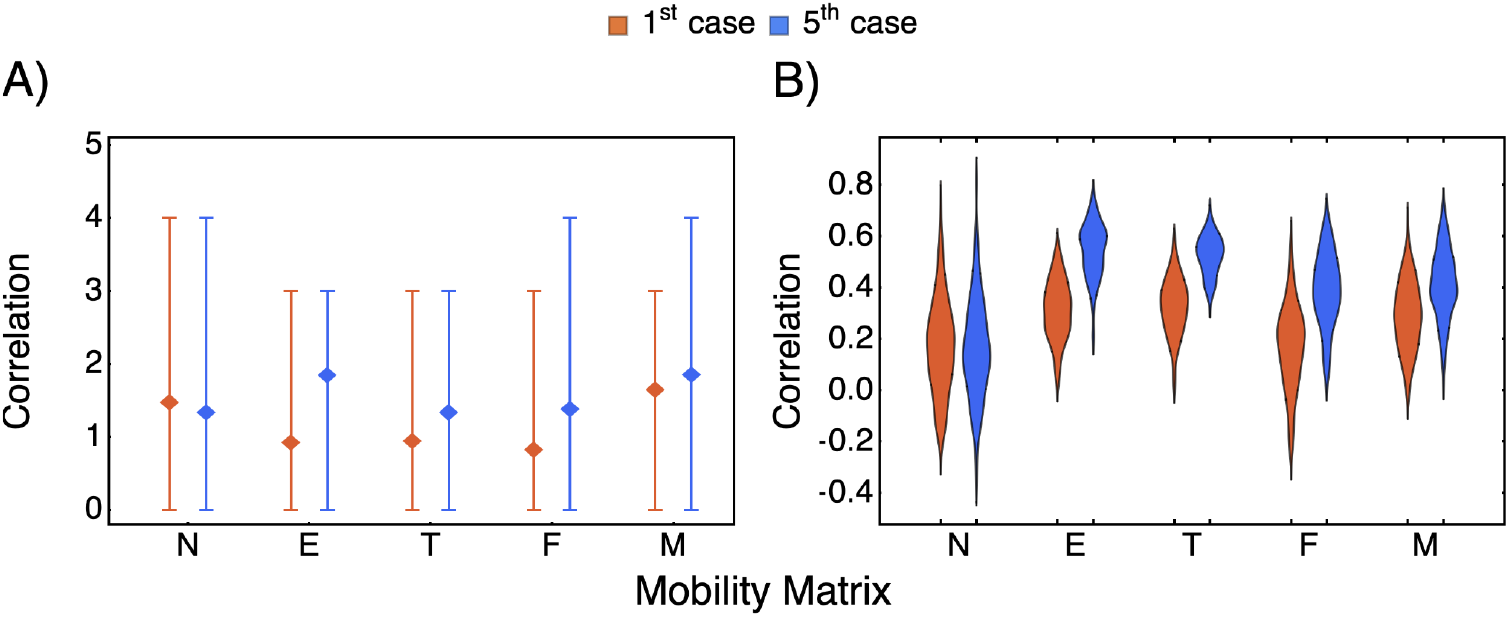
The simulated and reported ranks for various mobility matrices. A) The overlap (mean, minimum, and maximum number of regions correctly predicted) according to the first five regions reporting infection. B) The distribution of the Spearman rank correlation between each replicate and the reported rank. N, E, T, F, and M denote respectively the null model, the gravity model based on euclidean distance between centroids, the gravity model based on transit time, the Internal Migration Flow, and the mobile phone model, respectively. The reported ranks are either based on the 1^st^ (red) or the 5^th^ (blue) confirmed case (Analamanga is excluded as it was used as the initial condition).

In comparing simulated versus reported ranks, the overall median Spearman rank correlation was highest for the model using the gravity based on Euclidean distance (0.56), followed by the gravity based on transit time (0.53), the mobile phone model (0.4), and the Internal Migration Flow (0.38) (Fig. 4B). Increasing the exponent *τ* in the gravity matrix improved the predictive ability of the metapopulation model (Fig. S2 for *τ* = 0. 5, 1, 1. 5, the median of the distribution of the correlation increases from 0.45, 0.53, to 0.56 for the gravity based on Euclidean distance and from 0.44, 0.49, to 0.53 for gravity based on transit time). Interestingly, the highest correlation for the 5^th^ case among all replicates was with the null model with a value of 0.81.

Given the challenge in predicting the order of the reported cases, we investigated if some regions are more difficult to predict than others. Overall, the southern area of the country was consistently the most difficult to predict (Fig. 5). However, for all other areas of the country, there were no consistent patterns by the type of model.

**Figure 5:**
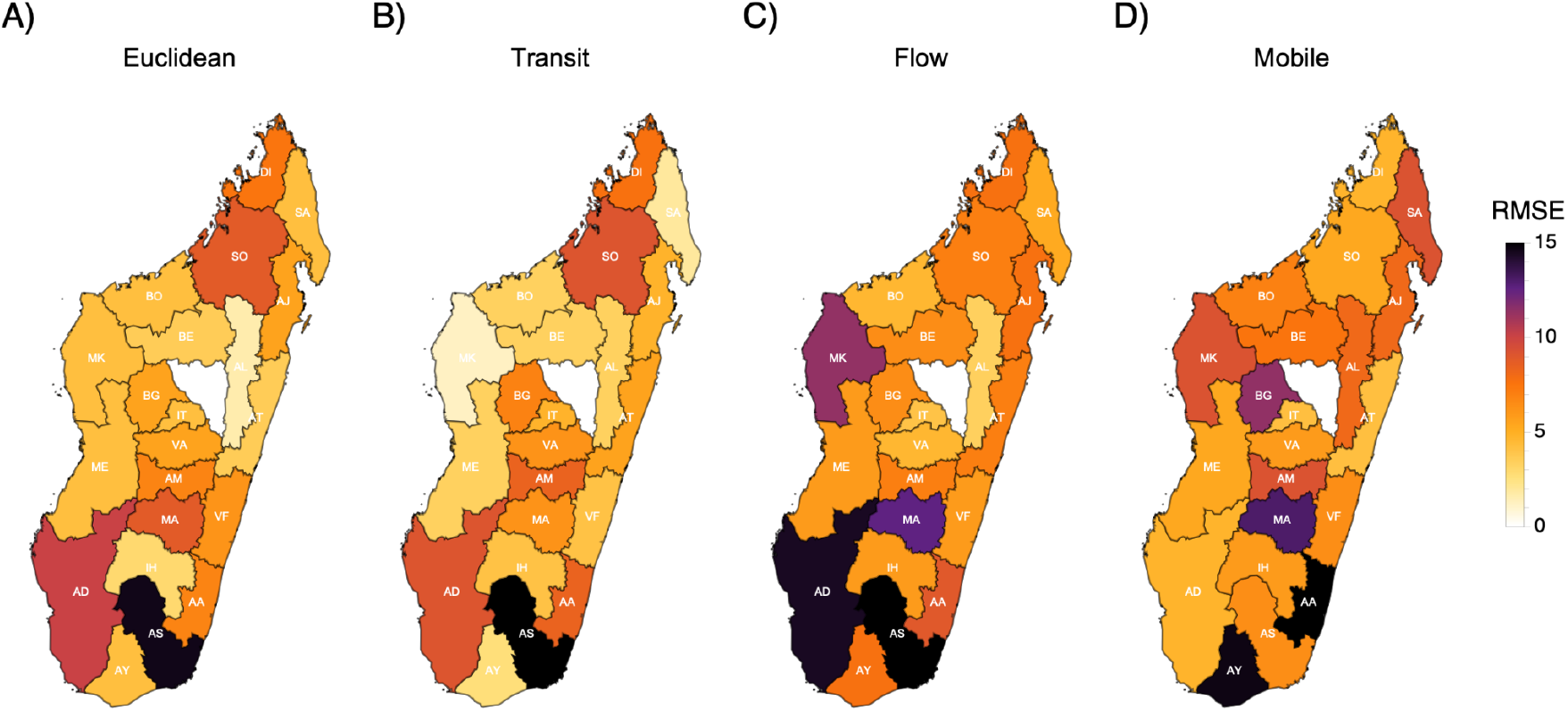
The root mean squared error (RMSE) in predicting the rank of the reported fifth case for each region. The regions are colored based on the accuracy of the ranks for the reported fifth case for each region using various mobility matrices: A) Euclidean, B) transit, C) Internal Migration Flow (Flow), and D) mobile phone data.

Finally, we compared the performance of each mobility matrix with a statistical approach. Table 1 shows the estimated parameter, *s*, representing the importance of the matrix in explaining the pattern, and the significance value. For the 1^st^ case, the models are quite similar showing intermediate s (0.5 < *s* < 0.8) but none are significantly different from a null model. For the 5^th^ case, the results are inconsistent with regards to the best mobility matrix compared to predictions of the mechanistic model. The gravity model based on transit performs poorly with the lowest s, while the Internal Migration Flow model performs best, but is not statistically significantly better than the null model. Overall, only the mobile phone model was significantly associated *(p<0*.*02)* with the order of detection, and only for the 5^th^ case. Complete results for all mobility matrices are shown in Table S1.

**Table 1:** The estimated s (p-value) of the network-based diffusion analyses for the four main mobility matrices used. Significant p-value in bold.

|  | Euclidean | Transit | Flow | Mobile Phone |
| --- | --- | --- | --- | --- |
| 1st case | 0.68 (0.19) | 0.51 (0.33) | 0.65 (0.37) | 0.82 (0.12) |
| 5th case | 0.64 (0.29) | 0.02 (0.98) | 0.99 (0.18) | **0.98 (0.02)** |

## Discussion

Understanding what types of mobility data and models can best predict spatial dynamics of infectious diseases, and particularly emergent pathogens, could importantly contribute to allocating scarce resources, prioritizing where improvements in healthcare and surveillance will be vital, and estimating the possible pace and severity of the epidemic (Grenfell et al., 2001; Rice et al., 2021; Tatem et al., 2006; Zhou et al., 2020). Often in low and middle income settings, there are few data sets on human travel available (Wesolowski et al., 2016), and limited surveillance data to estimate spatial dynamics directly from the pattern of cases (e.g., as in (Bjørnstad and Grenfell, 2008)). Here, we leverage a range of possible data sources on human mobility in order to better understand the spread of SARS-CoV-2, by integrating matrices describing mobility between regions into a spatial model of SARS-CoV-2 and a network-based diffusion analysis. By comparing the simulated trajectories with data, we evaluate the ability of different measures of mobility to predict the spatial spread of SARS-CoV-2 in Madagascar.

A major challenge in approaches of this kind is data availability. First, from the side of infectious disease data, there is limited availability of case numbers, and although many major cities in Madagascar have uniquely detailed mortality records (Masquelier et al., 2019) with scale and scope adequate to detect major outbreaks (Rasambainarivo et al., 2021) data compilation and accessibility to the research and public health community have lagged. To fill this gap in the landscape of public health communication in Madagascar, we developed a dashboard by collating data from daily televised reports, and this is the data that we use in our analyses. The quality of the data on cases can thus only be as good as these available reports. For instance, daily reporting was interrupted between the 13^th^ of October 2020 and the 13^th^ of March 2021 and was replaced by weekly cumulative numbers.

Uncertainties in the case data will be of most concern if there are marked spatial differences in reporting. Our analysis indicates large differences in the rank of the regions confirming the first case and the fifth case. The first case is most likely driven by imported cases, while the fifth case is likely to emerge as a result of onward transmission. Importantly, different locations may have different probabilities of both early detection, and onward transmission. As an example, Menabe and Diana were among the first five regions to report a first case but then lagged before the fifth case was reported. These regions represent popular tourist destinations and access by air is easy. Perhaps in part due to this demographic, the first cases in these regions were thus quickly isolated, contact tracing was swiftly established, and thus chains of transmission were slow to develop, delaying the fifth reported case. One positive interpretation of this pattern is that with adequate testing and contact tracing (as we hypothesize could have occurred following detection among tourists) spread from imported cases could be quickly controlled. Rolling out and prioritizing these strategies early on can thus have had an impact on curbing disease spread. Typically, data on air travellers is digitized and detailed, and could be leveraged to identify most regions at risk of such early introductions.

Moving from availability of case data to considering availability of mobility data, there are also a set of challenges at this end. The first three mobility matrices that we use were formulated from gravity-based models with varying degrees of realism, encompassing for example the distance between two regions calculated using Euclidean distance between the centroid of the regions or the actual transit time from transport companies; while the fourth mobility matrix we use is directly based on mobile phone data. Our comparison focussed on relative magnitudes of movement between regions rather than absolute magnitudes of movement, given the various uncertainties in the data available to develop a fully parameterized model. None of these approaches were able to correctly predict all of the reported patterns of spatial spread in Madagascar, although on average they all performed better than a null model. The best performing models were either the simplest (gravity with Euclidean distance) or extrapolated from data on mobility (mobile phone data). Adding realism in the gravity model with either transit data or Internal Migration Flows did not improve prediction. In a few rare replicates, the null model actually generated the most accurate predictions indicating the unpredictable nature of spread. Yet on average, the simplest gravity model most likely captures the core diffusion component of the process (infection ultimately spills into neighboring regions when the number of cases is high enough). Inference based on mobile phone data, despite being processed over five years ago (Ihantamalala et al., 2018; Wesolowski et al., 2016), had the best performance, perhaps because it captures a wider diversity of connections than are commonly predicted by a gravity model (Ihantamalala et al., 2018; Oliver et al., 2020; Tizzoni et al., 2014; Wesolowski et al., 2016, 2015a, 2015b). In fact, the simplest gravity and mobile phone models performed well in different (non-overlapping) regions, suggesting that they capture different important aspects of mobility.

Notably, some approaches performed strikingly poorly (e.g., the internal migration flow model) indicating a need for caution in deciding what model or metric of mobility to use. Finally, the performance of the mobile phone data relative to other measures strongly suggests that accurate up-to-date measurements of mobility from this source (rather than the 2015 data we were compelled to use) might have opened the way to anticipating spread and reacting appropriately. Designing regulatory pipelines that efficiently enable sharing of detailed yet anonymous mobility from mobile phone companies in such times of crisis should be a priority in Madagascar, as it has been elsewhere (Buckee et al., 2020; Grantz et al., 2020; Kishore et al., 2020; Oliver et al., 2020).

As most models did not reliably predict the rank of the timing of the first, nor fifth case per region (Figs. 4 and 5) this work is still some steps away from driving policy recommendations. In particular, all models largely failed to predict the spatial patterns in the South, less populated and connected area of the country, possibly because stochasticity and thus rare events are overwhelmingly important, and perhaps also because delays in testing and data reporting given the remoteness of the region blurred the signal in numbers of cases. A key direction for expanding this work is to identify where models and data provide reasonable predictions and where they do not. The analysis reported here provides a potential starting point for further sensitivity analyses that explores core drivers of expectations of outcomes given the topography of the network, providing general expectations for pathogen spread. Uncertainty in case data is a very general issue in developing a mechanistic understanding of infectious diseases (for example, case numbers often apparently paradoxically increase with vaccination coverage, but this is actually a result of concomitant improvements in surveillance (Prada et al., 2018)). Various approaches to correcting for biases are available (Becker and Grenfell, 2017; Jarvis et al., 2021), but transparency in data generation mechanisms is an essential component.

There are a number of caveats associated with this work. In particular, by focussing simply on region population size and connectivity patterns, the model simplifies a number of aspects that may be important to the pace of spread of SARS-CoV-2, such as within region dynamics (i.e., some regions may be more internally connected than others (Rice et al., 2021)), as well as interventions including travel bans and how these changed connectivity over the first months of spread. However, the better performance of the mobile phone model compared to the gravity-based model in both the mechanistic and statistical model suggests that the connectivity matrix used to link the regions is the core of the problem. Our analysis provides a first step for moving towards models that can capture the spread of an emergent pathogen. It also highlights the centrality of data availability and strengthening collaboration among different institutions with access to critical data - models are only as good as the data that they use - so building towards effective data-sharing pipelines is essential.

## Data Availability

Data available by request.

https://github.com/ramiadantsoa/mobilityMada

## Authors Contributions

TR, BLR, CJEM, AW, FR conceived and designed the paper, TR and FR wrote and performed the analyses. SR gathered the data from the bus company, TR, BLR, CJEM, AW, FR, AHR, and FMR wrote the manuscript.

The authors declare no competing interests

## Acknowledgements

We thank Orange for sharing the original mobile phone data. We also thank Valerie Ranaivoson, Domoina Nadia Andriamamenosoa and Faly Aritiana Fabien for their assistance, support, and encouragement.

## Supplements

Full description of the standardization of the mobility matrix.

In raw format, the mobility matrix is a table with 22 rows and 22 columns representing pairwise mobility which in principle can take any values. For instance, the raw mobile phone data is based on monthly movement and the gravity matrix can be scaled as desired by changing the scalar *a* in the main text. The mobility is divided into three components: the number of individuals leaving each region (X(i), *i* is for region), the infection status of these individuals (X_K_(i)) and how they are distributed to the other regions. First we set the diagonal of the raw matrix to zero as we are not considering within-region mobility. The number of individuals moving out of a region i is 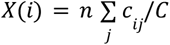 where 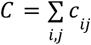 where n is a scaling factor and is the same for all regions. The number of individuals moving per status is drawn from a multinomial distribution *X*_*K*_(*i, t*) ∼ Multinomial *(X(i), pS(i, t), pE(i, t), pI(i, t), pR(i, t))* where *pS, pE, pI*, and *pR* are the proportion of individuals in each infection status. Finally, we converted each entry of the mobility matrix to probability, i.e., the connectivity *c*_*ij*_ becomes *p*_*ij*_ where 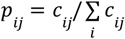 with *c*_*ii*_ = 0. These probabilities become the parameters of the multinomial distribution. The number of individuals moving from region *i* to *j* and is drawn from a multinomial distribution: Multinomial (*X*_*K*_(*i*), *p*_*i*1_, … *p*_*ij*_ … *p*_*i*22_).

**Table S1:** The estimated s (p-value) of the network-based diffusion analyses for all eight mobility matrices used. TThe numbers 1, 2, and 3 refer respectively to *τ* = 0.5, 1, 1.5, the exponent in the numerator term in the gravity model. Significant p-value in bold.

|  | Euclidean 1 | Euclidean 2 | Euclidean 3 | Flow | Transit 1 | Transit 2 | Transit 3 | Mobile phone |
| --- | --- | --- | --- | --- | --- | --- | --- | --- |
| 1st case | 0.61 (0.31) | 0.68 (0.21) | 0.68 (0.19) | 0.65 (0.37) | 0.49 (0.32) | 0.52 (0.31) | 0.51 (0.33) | 0.82 (0.12) |
| 5th case | 0.75 (0.27) | 0.71 (0.27) | 0.64 (0.29) | 0.99 (0.18) | 7.83e-5 (1) | 7.83e-5 (1) | 0.02 (0.98) | **0.98 (0.02)** |

## Supplementary figures

**Fig S1:**
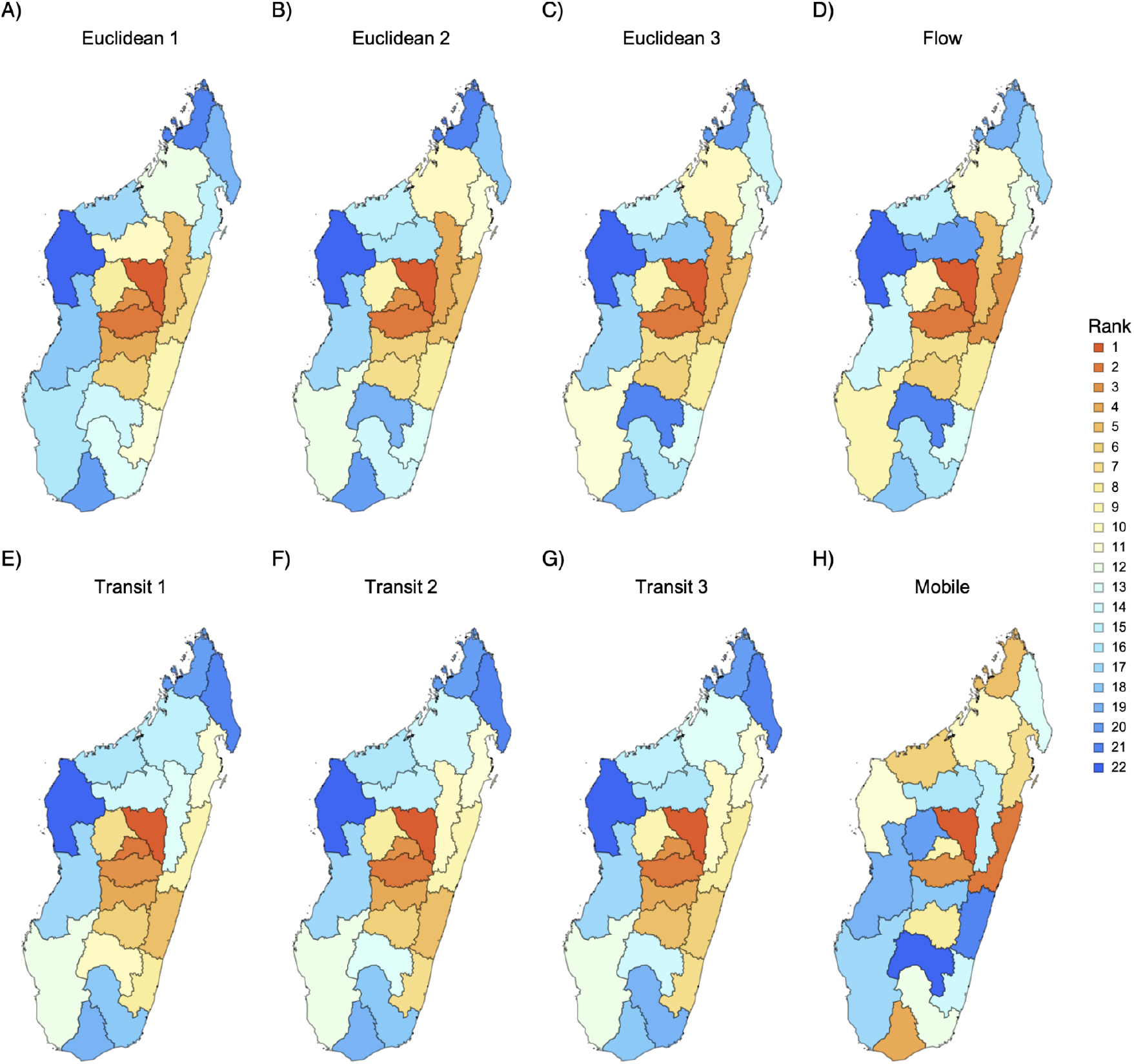
Comparison of the all mobility matrices used throughout. The regions are ranked according to the number of individuals traveling to that region per day. Euclidean (A-C) and transit (E-F) mobility matrices constructed from the gravity based model where distance is either based on euclidean distance between centroid or transit time, respectively. The numbers 1, 2, and 3 refer respectively to *τ* = 0.5, 1, 1.5, the exponent in the numerator term in the gravity model. Flow (D) is short for Internal Migration Flow--a gravity based model downloaded from worldpop.org. Mobile (H) represents mobility matrix inferred from mobile phone data.

**Fig S2:**
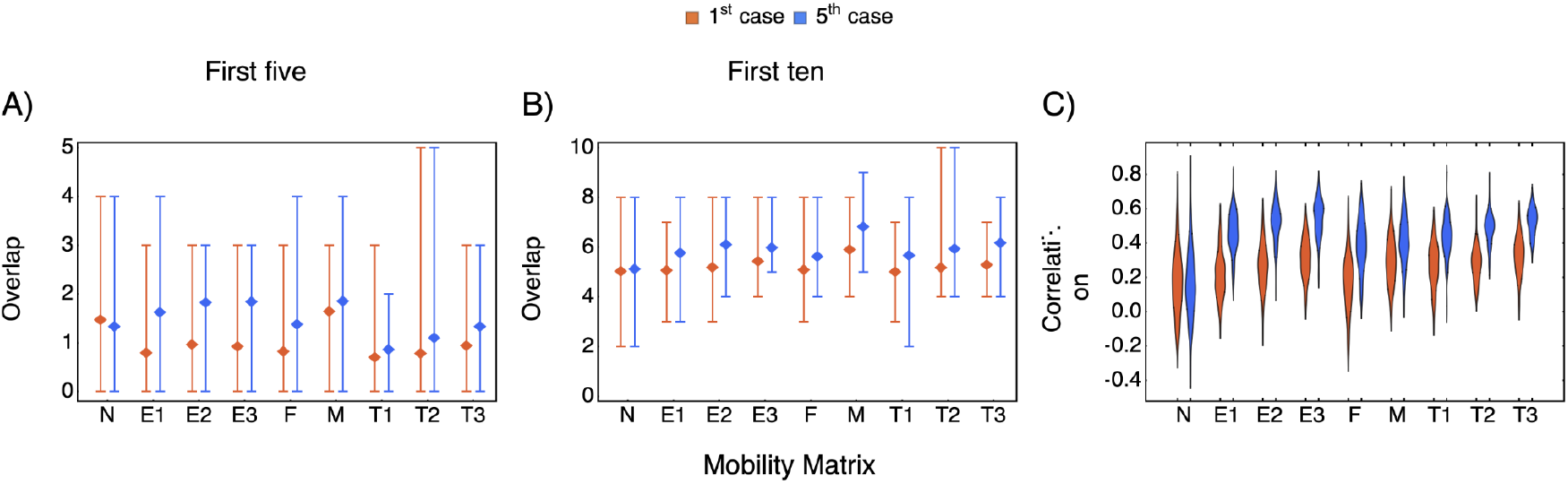
Comparing empirical and simulated ranks for all mobility matrices considered. A) and B) The overlap (mean, minimum, and maximum number of regions predicted) according to the first five and first ten regions with infection, respectively. C) The distribution of the Spearman rank correlation between the simulated and the true rank. On the x-axis, the true patterns are either based on the first or first fifth confirmed case (Analamanga is excluded as it was used as the initial condition). N, I, and O denote respectively the null model, the Internal Migration Flow, and the matrix derived from the mobile phone data. E and T represent mobility matrices derived from the gravity based model where distance is either based on euclidean distance between centroid or transit time. The numbers 1, 2, and 3 refer respectively to *τ* = 0.5, 1, 1.5, the exponent in the numerator term in the gravity model. Number of replicates: 100.

**Figure S3:**
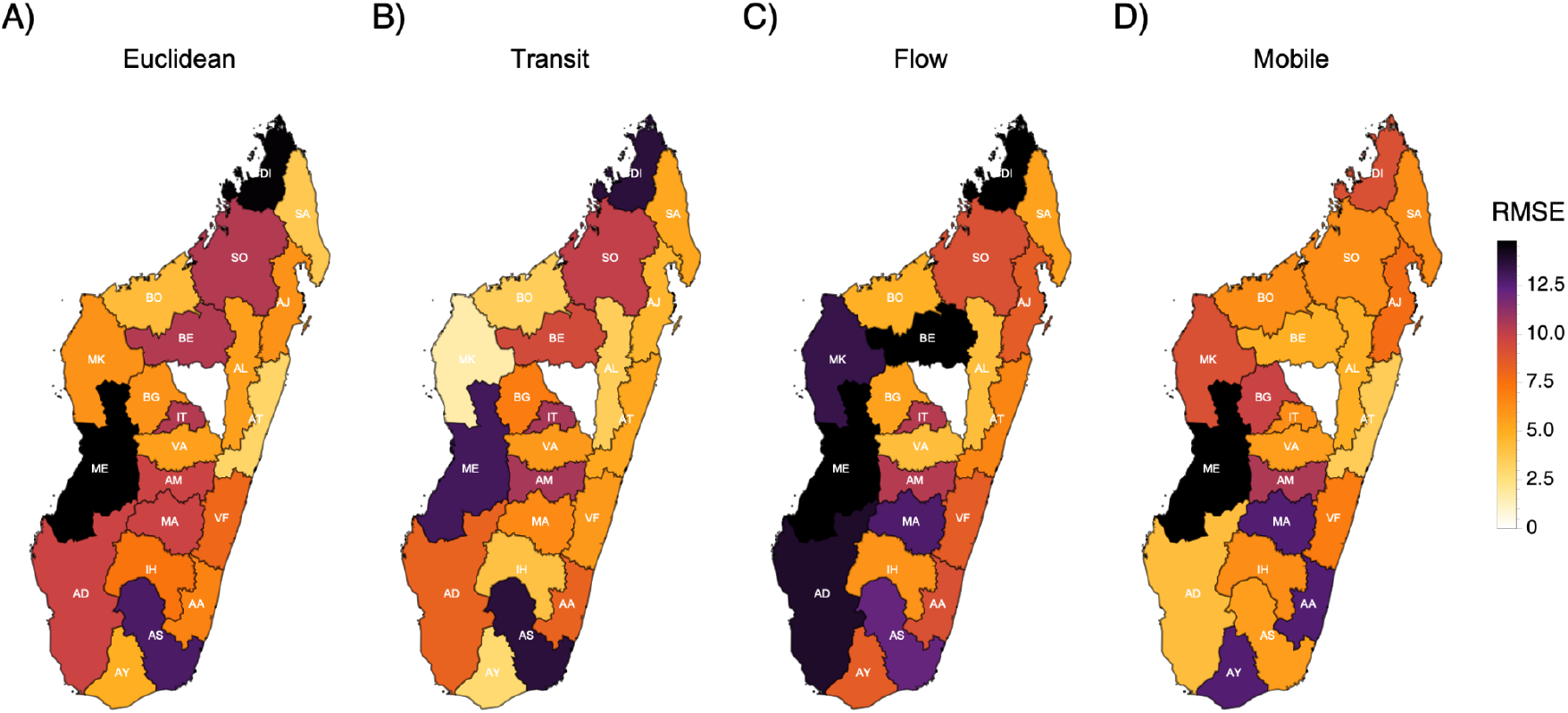
Root mean squared error in predicting the rank of the first case for each region. Euclidean (A) and transit (B) are gravity based models where distance is either based on euclidean distance between centroid or transit time by bus, respectively. Flow (C) is short for Internal Migration Flow and is a gravity based model downloaded from worldpop.org. Mobile (D) represents mobility matrix inferred from mobile phone data.

